# High-frequency EEG activity moderates the association between amyloid burden and tau in cognitively normal adults

**DOI:** 10.64898/2026.10.30.26366850

**Authors:** Seyed Hani Hojjati, Ana Radanovic, Isabelle Martin, Ludvik Alkhoury, Tracy H Butler, Nancy Foldi, Sudhin A Shah, Gloria C Chiang

## Abstract

**Background:** Resting electroencephalogram (EEG) may capture early excitatory-inhibitory (E-I) imbalance in Alzheimer’s disease, but its relationship to beta-amyloid (Aβ) and tau in cognitively normal adults is unclear.

**Methods:** We analyzed resting-state EEG from 24 older adults: 21 cognitively normal (13 Aβ-negative, 8 Aβ-positive) and 3 with MCI/AD (all Aβ-positive), with Aβ positron emission tomography (PET); 20 cognitively normal adults also had tau PET. Four parietal EEG measures reflecting excitation-inhibition (E-I) balance and neuronal excitability (periodic gamma power, alpha-gamma and theta-gamma phase-amplitude coupling [PAC], and the aperiodic exponent) were assessed for associations with Aβ burden and early medial temporal lobe (MTL) tau, adjusting for age and sex.

**Results:** In 21 cognitively normal adults, higher Aβ burden was associated with higher parietal periodic gamma power (β = +0.50, p = 0.025). No E-I measure was associated with MTL tau across all participants. However, in Aβ-positive adults, higher periodic gamma was associated with higher MTL tau, most strongly in the entorhinal cortex (ρ = +0.90, p = 0.002), with no association in Aβ-negative adults. Importantly, periodic gamma also moderated the association between Aβ and MTL tau (p = 0.012; consistent across MTL regions after FDR correction).

**Conclusions:** In this pilot study, parietal periodic gamma, an EEG marker of cortical excitability, increased with Aβ burden and was related to MTL tau only in the presence of Aβ. Periodic gamma power may serve as a candidate marker of Aβ-associated E-I imbalance and early AD progression.

## 1. INTRODUCTION

Beta-amyloid (Aβ) deposition is a core pathological hallmark of Alzheimer’s disease (AD) that can be detected decades before the onset of clinical symptoms, providing a substantial window of opportunity for potential therapeutic interventions.^1, 2^ A multinational study of over 6000 participants reported that Aβ-positive individuals had only a 17% risk of progressing to mild cognitive impairment (MCI) or AD within 5 years.^3^ Despite our increased ability to detect Aβ pathology early, with Aβ deposition present in approximately 20-30% of cognitively normal older individuals^4^, there is an urgent clinical need to identify which individuals will actually develop clinical AD in the near term^5^. This knowledge gap raises concerns for potential overdiagnosis and overtreatment of Aβ-positive individuals, without clear identification of those most likely to benefit from targeted therapies, resulting in the 2024 International Working Group recommending incorporating cognitive measures into disease management frameworks^3^.

Aβ is considered the initiator of a sequence of events that lead to neuronal degeneration, memory decline, and subsequent AD.^6^ Its role as a Core 1 biomarker of AD underlies the recently proposed National Institute on Aging-Alzheimer’s Association “ATN” framework^7^ and revised criteria^8^. However, the mechanisms by which early Aβ pathology disrupts brain function, leading to tau elevation and disease progression, remain unclear. Aβ does not accumulate uniformly across the cortex. Previous studies show that Aβ first deposits in the posterior medial and lateral parietal cortex, particularly the precuneus and posterior cingulate, which are core hubs of the default mode network (DMN).^9, 10^ These hubs have the highest lifelong levels of neural activity and metabolism in the brain.^11^ We and other resting-state functional magnetic resonance imaging (rs-fMRI) studies^12-14^ have shown that early network reorganization (i.e., hyperconnectivity) occurs in response to abnormal Aβ in presymptomatic stages of AD. We provide unique preliminary evidence that the limbic-DMN network exhibits Aβ-related aberrant network hyperconnectivity, involving key memory- and tau-related limbic regions^15-18^. We speculated that this aberrant network hyperconnectivity is due to Aβ-related excitatory-inhibitory (E-I) imbalance and may drive the tau accumulation and disease progression. Whether this Aβ-related E-I imbalance can serve as an early predictor of tau spread and disease progression remains to be demonstrated.

These knowledge gaps limit our ability to use mechanistically grounded markers for patient stratification and early targeted intervention before irreversible neurodegeneration occurs. Together, these findings support a hypothetical model in which early Aβ-driven disruption of E-I balance manifests as detectable cortical hyperexcitability and tau accumulation and spread at early stages. In this study, we used a panel of electroencephalogram (EEG)-based measures to test whether EEG-based indices of E-I balance moderate the association between Aβ burden and early tau accumulation. This study may provide preliminary results for non-invasive EEG-based biomarkers of network dysfunction that precede tau spread in cognitively normal adults, offering a critical window for therapeutic intervention before widespread neurodegeneration occurs.

## 2. METHODS

### 2.1. Participants

We included 24 participants: 21 healthy controls (HC) for the main analysis, and 3 with mild cognitive impairment or early AD for further validation of E-I imbalance, Aβ burden, and early tau accumulation. All individuals underwent resting-state EEG, Aβ-PET imaging (18F-florbetaben or Pittsburgh Compound B [PiB] tracers), MRI (3D T1-weighted), and cognitive assessment; 20 HC individuals also underwent FDA-approved 18F-MK6240 tau-PET. The study was approved by the Institutional Review Board of Weill Cornell Medicine, and all participants provided written informed consent.

### 2.2. Neuroimage acquisition protocols

#### 2.2.1. MR acquisition

MR imaging was performed using a high-resolution 3D T1-weighted sequence (TR/TE = 2400/2.96 ms, flip angle = 9°; FOV = 25.6 × 25.6 cm, matrix size = 256 × 256, and 208 sagittal slices).

#### 2.2.2. Aβ-PET acquisition

Participants received either the 18F-florbetaben or 11C-PiB tracer via an intravenous catheter in the arm. For 18F-florbetaben, a dose of 300 MBq (8.1 mCi) ± 10% was administered as a single IV bolus, and brain images were acquired starting 90 minutes after injection, with four 5-minute frames (20 minutes). For 11C-PiB, a dose of 555 MBq (15 mCi) ± 10% was administered as a single IV bolus, and brain images were acquired starting 50 minutes after injection, with four 5-minute frames (20 minutes).

#### 2.2.3. Tau-PET

Participants received the 18F-MK6240 tracer via an intravenous catheter in the arm, with an injection of 185 MBq (5 mCi) ± 20% (maximum volume 10 mL), administered as a single IV bolus within 60 seconds or less (equivalent to 6 sec/mL max). Brain images were acquired starting 40 minutes after tracer injection, with four 5-minute frames (20 minutes).

#### 2.2.4. EEG acquisition

EEG data were recorded using a high-density 128-electrode saline-based EEG system (GSN-Hydrocel 128, Magstim-EGI, Eugene, OR, USA) based at the BHII/WCM. Electrode impedances were maintained below 50 kΩ throughout the recording session, and signals were referenced to the Cz electrode during acquisition. Each subject underwent 2 minutes of eyes-open resting-state EEG recording to capture resting neural activity.

### 2.3. MR and PET quantification process

#### 2.3.1. Structural MRI

The sMRI scans acquired on 3T scanners were processed using the FreeSurfer segmentation and cortical parcellation software package.

#### 2.3.2. Aβ-PET processing

Participants received PIB or FBB tracers. Images have been processed using our Centiloid pipeline^19^, in concordance with the Global Alzheimer’s Association Interactive Network (GAAIN) framework, which was developed to harmonize Aβ-PET quantification across tracers.^20^ Global Aβ burden was quantified as Centiloid. Aβ positivity was defined a priori as Centiloid>18.

#### 2.3.3. Tau-PET processing

We calculated Tau-PET standardized uptake value ratio (SUVR) from summed images over specific time intervals (40-60 min)^21^, using the cerebellar cortex as the reference region. Regional tau burden was quantified as SUVR within a FreeSurfer-based parcellation, with each region eroded by 1 voxel to minimize partial-volume effects. Analyses were restricted a priori to medial temporal lobe (MTL) targets: entorhinal cortex, hippocampus, amygdala, parahippocampal cortex, and an MTL composite. Bilateral subregional values were computed as the arithmetic mean of left and right hemisphere SUVR.

### 2.4. EEG quantification process and measures

#### 2.4.1. EEG processing

All preprocessing was performed via custom scripts using the MNE-Python software package.^22^ Raw EEG data were first re-referenced to the average of the bilateral mastoid electrodes. A notch filter was applied at 60 Hz and its harmonics using MNE-Python’s `notch filter` function. The filter was implemented as a zero-phase FIR (finite impulse response) filter with a Hamming window, a 1 Hz notch width, and a 10 Hz transition bandwidth. Data were then bandpass filtered between 1 and 120 Hz using MNE-Python’s default zero-phase FIR filter settings. We visually inspected the data to mark recording segments for removal and channels with excessive noise. Independent Component Analysis (ICA) was applied to remove artifacts, with visually identified eye blink and cardiac components excluded. ICA was performed using the PICARD algorithm.^23^ Bad channels were then interpolated using spherical spline interpolation. 90 seconds of the 2 minutes of eyes-open resting-state data were segmented into non-overlapping 3-second epochs for subsequent analysis. Automated epoch rejection was performed using the Autoreject.^24^ The data were downsampled to 250 Hz. We considered only channels restricted a priori to the 9-channel parietal region (P1-P8, Pz), given the spatial relevance to Aβ pathology and our hypothesis that Aβ-related hyperexcitability in default mode network hubs, detectable as altered E-I balance in parietal EEG, is associated with tau accumulation in the MTL.

#### 2.4.2. Spectral measures, spectral parameterization, and phase-amplitude coupling

Because our hypothesis targets cortical hyperexcitability, the primary analyses used a panel of four measures related to E-I balance and excitability: parietal periodic gamma power, alpha-gamma and theta-gamma phase-amplitude coupling (PAC), and the aperiodic exponent, a broadband index of E-I balance.

The FOOOF algorithm (version 1.1.1) was used to parameterize neural power spectra. Settings for the algorithm were set as: peak width limits: 2-20Hz; max number of peaks: 6; minimum peak height: 0.10; peak threshold: 2; and aperiodic mode: knee. Power spectra were parameterized across the frequency range 2 to 45Hz. Periodic (oscillatory) power in a given band was computed as the linear-power difference between the model’s full fitted spectrum and its aperiodic component alone (i.e., the peak component in isolation), rectangular-integrated within band edges (periodic gamma: 30-45 Hz) and averaged across parietal channels; the aperiodic exponent and offset were taken directly from each channel’s fitted aperiodic parameter in gamma frequency and averaged across parietal channels. Model goodness-of-fit was assessed for every channel fit using r-squared and the mean absolute error (MAE) between the model and the power spectrum. Across the 189 parietal channel fits in the 21 HC, the median r-squared was 0.98 (range 0.41-0.99; individual-level mean r-squared 0.79-0.99) and the median MAE was 0.050 (range 0.023-0.134, log10 power units). Fits with r-squared<0.90 were rare (11 of 189 channel fits). Due to the limited sample size of this pilot study and acceptable goodness-of-fit metrics, no participants were excluded.

For phase-amplitude coupling (PAC), phase- and amplitude-providing signals were extracted per channel with a 4th-order zero-phase (forward-backward) Butterworth bandpass filter (phase: alpha, 8-13 Hz; amplitude: gamma, 30-45 Hz), and instantaneous phase and amplitude were obtained via the Hilbert transform. The amplitude envelope was binned into 18 phase bins; the modulation index is the Kullback-Leibler divergence of the resulting phase-binned amplitude distribution from a uniform distribution, normalized by log(18) so that MI=0 indicates no coupling. Only within-channel (same-channel phase and amplitude) coupling was computed, then averaged across the 9 parietal channels. Raw modulation indices are reported as the primary PAC measure. Theta-gamma PAC was computed in the same way with theta (4-8 Hz) as the phase-providing band.

### 2.5. Statistical analysis

All continuous models used ordinary least squares with age and sex as covariates; effect sizes are partial correlations from the model t-statistic. Analyses were each FDR-corrected (Benjamini-Hochberg) within its own sections. Measures with a skewed distribution (absolute skewness > 2) were log10-transformed to limit the influence of extreme values. The robustness of key associations was assessed with heteroskedasticity-consistent (HC3) standard errors, Huber robust regression, and rank-based models.

#### 2.5.1. Aβ-EEG relationship

For the Aβ-EEG relationship, the primary analysis tested the continuous association between Centiloid and each of the four E-I measures within the 21 HC participants (measure ∼ Centiloid + age + sex, with measure and Centiloid z-scored so that β is standardized), FDR-corrected across the four EEG measures. The three groups (HC Aβ-negative, HC Aβ-positive, MCI/AD Aβ-positive) were also compared with one-way ANOVA, Welch-type tests, and Tukey’s HSD.

#### 2.5.2. EEG-tau relationship

The four E-I measures were tested against tau in the MTL composite (primary target) and in four MTL subregions (entorhinal, hippocampus, amygdala, parahippocampal) in the 20 HC participants with tau-PET. A direct association model (tau ∼ EEG + age + sex) was tested first. To examine whether the relationship depended on Aβ, the same associations were tested separately in Aβ-positive and Aβ-negative HC; because of the small group sizes, these within-group analyses used correlations without covariates, with Spearman’s rank correlation as the primary statistic in the Aβ-positive group and Pearson’s correlation also reported.

Then, we tested whether Aβ moderated the EEG-tau association with the model tau ∼ EEG × Centiloid + age + sex, on the same measures and regions.

## 3. RESULTS

### 3.1. Cohort characteristics

24 participants were included: 13 HC, Aβ-negative (mean age 70.9 ± 8.1 years; 7 female; mean Centiloid -5.7 ± 6.7); 8 HC, Aβ-positive (mean age 74.9 ± 6.5 years; 5 female; mean Centiloid 52.5 ± 22.0); and 3 MCI/AD, all Aβ-positive (mean age 72.9 ± 13.0 years; 1 female; mean Centiloid 105.7 ± 47.1). All HC participants had a clinical dementia rating (CDR) of 0, and participants with MCI/AD had a CDR of 0.5 or 1. Full cohort characteristics are given in **Table 1**.

**Table 1.**
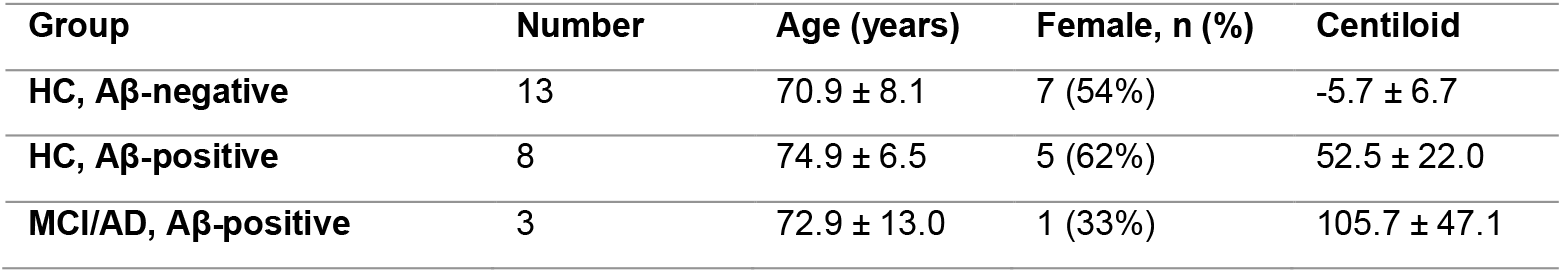
Cohort demographic and clinical characteristics.

### 3.2. Resting-state spectra in parietal cortex

Resting-state power spectra for parietal cortex are shown in **Figure 1**, averaged within each of the three Aβ/diagnosis groups. **Figure 1** shows that the resting-state parietal power spectra differed by Aβ and diagnostic group. In Aβ-positive adults, the peak was 3.7 dB (at 8 Hz), with lower theta and alpha power. In MCI/AD, the alpha peak was at 8 Hz and weakened to 2.0 dB, with higher low-frequency power and a flat high-frequency plateau about 10 dB above that of Aβ− adults at 30-45 Hz. Cognitively normal Aβ-negative and Aβ-positive adults already differ detectably, whereas the MCI/AD difference is much larger, with a rise in both low- and high-frequency power with a completely different pattern.

**Figure 1.**
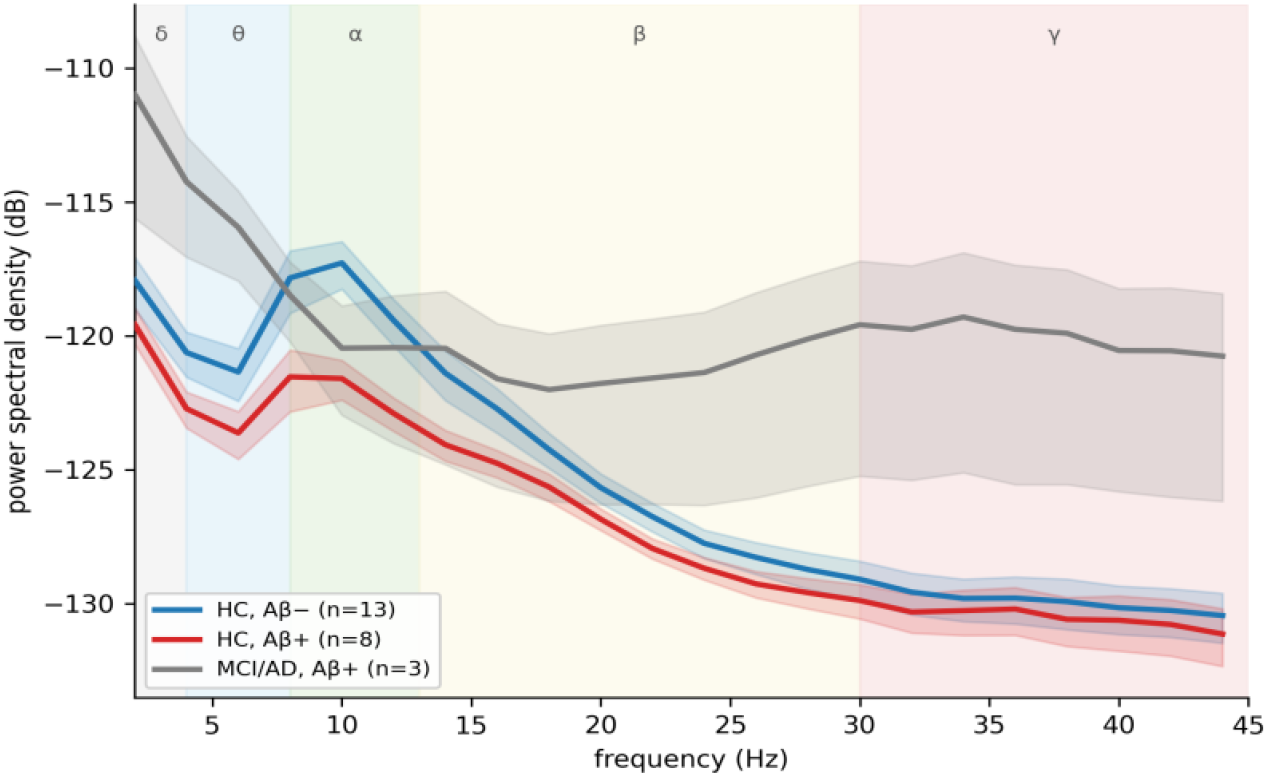
Resting-state power spectra in parietal cortex, by Aβ/diagnosis group (HC Aβ-negative n=13, HC Aβ-positive n=8, MCI/AD n=3).

### 3.3. Relationship between Aβ and parietal E-I measures

Within the 21 HC participants, higher Centiloid was associated with higher parietal periodic gamma (log10; β=+0.50, 95% CI 0.07 to 0.93, p=0.025; **Figure 2A, Table 2**). This association was consistent across estimators (HC3 p=0.017; Huber robust regression p=0.011; partial Spearman ρ=+0.53, p=0.014), although it did not survive FDR correction across the four E-I measures (q=0.098). Theta-gamma PAC (β=+0.38, p=0.11), alpha-gamma PAC (β=+0.29, p=0.23) and the aperiodic exponent (β=-0.18, p=0.47) were not associated with Centiloid.

**Table 2.** Association between Centiloid and parietal E-I measures in 21 HC participants (age- and sex-adjusted; standardized β; FDR across the four measures).

| **Measure** | **β (95% CI)** | **Partial r** | **p** | **q (FDR)** | **Partial Spearman ρ (p)** |
|---|---|---|---|---|---|
| **periodic gamma (log10)** | +0.50 (0.07, 0.93) | +0.51 | 0.025 | 0.098 | +0.53 (0.014) |
| **alpha-gamma PAC (log10)** | +0.29 (-0.20, 0.77) | +0.29 | 0.23 | 0.31 | -0.02 (0.92) |
| **theta-gamma PAC (log10)** | +0.38 (-0.09, 0.86) | +0.38 | 0.11 | 0.22 | +0.14 (0.54) |
| **aperiodic exponent** | -0.18 (-0.71, 0.34) | -0.18 | 0.47 | 0.47 | -0.02 (0.94) |

**Figure 2.**
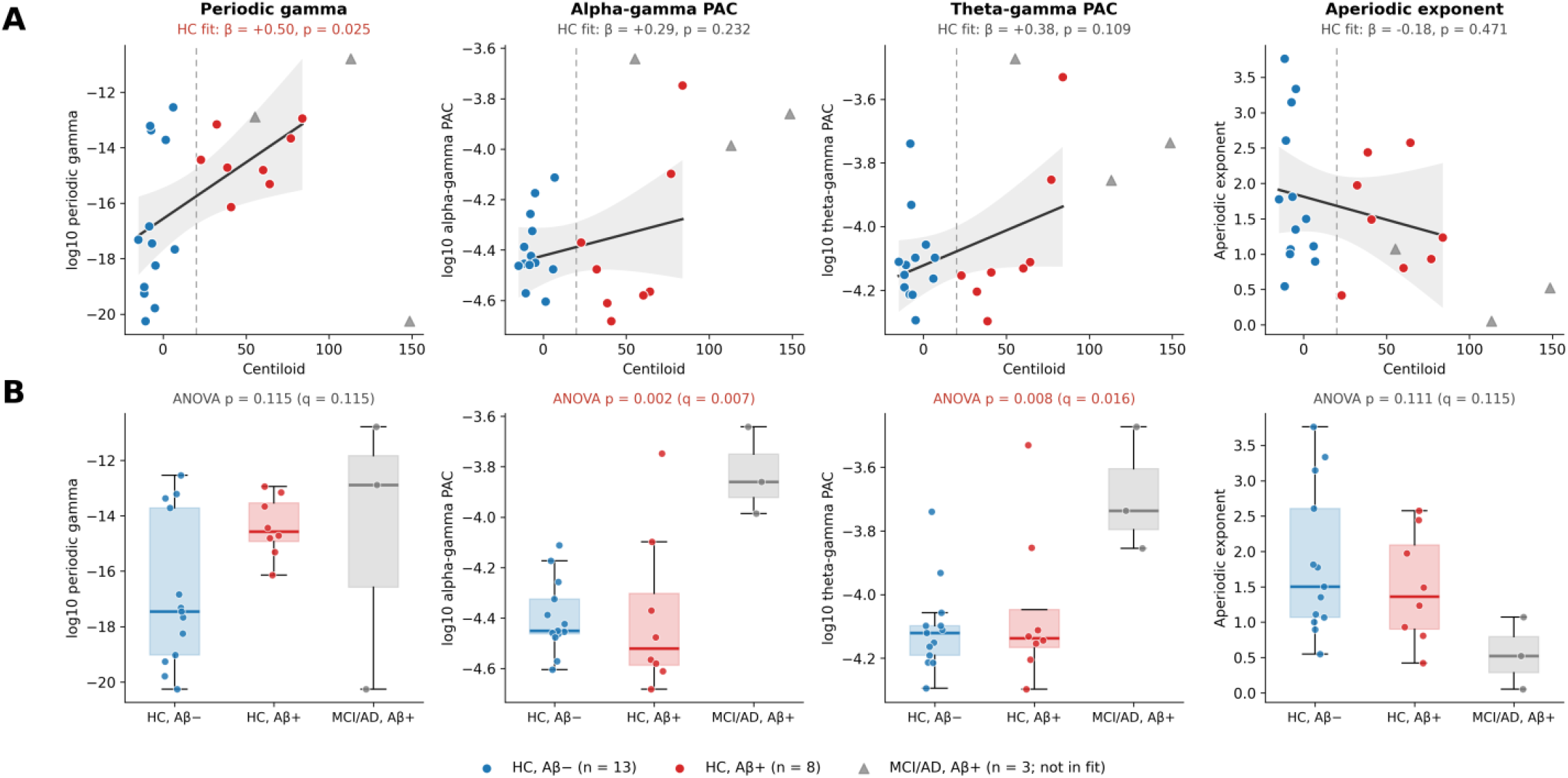
Parietal E-I measures and Aβ. (A) Association with Centiloid in the 21 HC participants (Aβ-negative, blue; Aβ-positive, red; MCI/AD shown as grey triangles for reference and not included in the fit). Lines and bands show the linear fit and 95% CI; β and p are from age- and sex-adjusted models. (B) Measures by Aβ/diagnosis group. Periodic gamma and PAC are log10-transformed. The significant associations are shown in red font.

Across the three groups, the two PAC measures differed (alpha-gamma ANOVA p=0.0018, theta-gamma p=0.0081), driven by higher values in MCI/AD (Tukey HC Aβ-negative vs. MCI/AD p=0.0017 and p=0.006), whereas Aβ-positive and Aβ-negative HC did not differ on any E-I measure (Tukey p>0.12; **Figure 2B**). Based on **Figures 1 and 2**, the MCI/AD pattern likely reflects changes associated with the clinical stage of disease.

Because our primary hypothesis focuses on Aβ-related E-I imbalance during the preclinical stage, all subsequent analyses were restricted to cognitively normal participants.

### 3.4. E-I measures and MTL tau: dependence on Aβ positivity

Across all 20 HCs with tau-PET (one HC individual did not have tau-PET), no E-I measure was associated with MTL composite or subregional tau after FDR correction (all q≥0.17; **Table 3**). The strongest nominal association was for alpha-gamma PAC and entorhinal tau (partial r=0.60, uncorrected p=0.0085), but it was not supported by rank-based correlation. However, the pooled analysis combines participants with and without Aβ. In the 8 Aβ-positive HC, higher periodic gamma was associated with higher tau in the MTL composite (ρ=+0.71, p=0.047) and in every subregion except the amygdala, most strongly in the entorhinal cortex (r=+0.79, p=0.020; Spearman ρ=+0.90, p=0.002; FDR q=0.040; **Figure 3**) and the parahippocampal cortex (ρ=+0.86, p=0.0065, q=0.065). The entorhinal association remained when any single participant was removed (r≥0.73). There was no such association in the 12 Aβ-negative HCs (entorhinal r=+0.09, p=0.78). The PAC measures showed nominal Pearson correlations in Aβ-positive HCs that were not supported by rank-based correlations, and the aperiodic exponent was unrelated to tau in either group. Thus, only periodic gamma was related to MTL tau in the presence of Aβ.

**Table 3.** Parietal E-I measures and MTL tau in 20 HC participants: direct association in all HC (age- and sex-adjusted partial r), Pearson (r) and Spearman (ρ) correlations within Aβ-positive (n = 8) and Aβ-negative (n = 12) HC, and moderation by Centiloid (age- and sex-adjusted). q: FDR across 20 tests within each family.

| Measure | Region | All HC: partial r (q) | Aβ+: r (p) | Aβ+: ρ (p; q) | Aβ−: r (p) | Aβ−: ρ (p) |
|---|---|---|---|---|---|---|
| **periodic gamma (log10)** | MTL composite | +0.08 (0.94) | +0.67 (0.068) | +0.71 (0.047; 0.19) | -0.14 (0.67) | -0.10 (0.76) |
| **periodic gamma (log10)** | entorhinal | +0.23 (0.63) | +0.79 (0.020) | +0.90 (0.002; 0.040) | +0.09 (0.78) | +0.01 (0.98) |
| **periodic gamma (log10)** | hippocampus | -0.05 (0.94) | +0.66 (0.073) | +0.71 (0.047; 0.19) | -0.22 (0.49) | -0.13 (0.68) |
| **periodic gamma (log10)** | amygdala | +0.04 (0.94) | +0.54 (0.17) | +0.55 (0.16; 0.46) | -0.25 (0.44) | -0.20 (0.53) |
| **periodic gamma (log10)** | parahippocampal | +0.22 (0.63) | +0.69 (0.060) | +0.86 (0.0065; 0.065) | +0.10 (0.75) | +0.09 (0.78) |
| **alpha-gamma PAC (log10)** | MTL composite | +0.51 (0.20) | +0.77 (0.025) | +0.45 (0.26; 0.58) | +0.07 (0.83) | +0.01 (0.97) |
| **alpha-gamma PAC (log10)** | entorhinal | +0.60 (0.17) | +0.85 (0.0082) | +0.74 (0.037; 0.19) | +0.22 (0.49) | +0.13 (0.70) |
| **alpha-gamma PAC (log10)** | hippocampus | +0.44 (0.20) | +0.73 (0.041) | +0.45 (0.26; 0.58) | +0.08 (0.80) | -0.06 (0.85) |
| **alpha-gamma PAC (log10)** | amygdala | +0.47 (0.20) | +0.69 (0.059) | +0.36 (0.39; 0.77) | +0.04 (0.91) | +0.05 (0.88) |
| **alpha-gamma PAC (log10)** | parahippocampal | +0.54 (0.20) | +0.82 (0.014) | +0.67 (0.071; 0.24) | +0.00 (0.99) | -0.06 (0.86) |
| **theta-gamma PAC (log10)** | MTL composite | +0.37 (0.28) | +0.69 (0.059) | -0.17 (0.69; 1.00) | -0.08 (0.80) | -0.03 (0.91) |
| **theta-gamma PAC (log10)** | entorhinal | +0.44 (0.20) | +0.72 (0.045) | +0.14 (0.74; 1.00) | +0.06 (0.85) | +0.06 (0.86) |
| **theta-gamma PAC (log10)** | hippocampus | +0.27 (0.56) | +0.62 (0.098) | -0.17 (0.69; 1.00) | -0.10 (0.76) | +0.03 (0.93) |
| **theta-gamma PAC (log10)** | amygdala | +0.38 (0.28) | +0.66 (0.077) | -0.12 (0.78; 1.00) | -0.16 (0.61) | +0.04 (0.90) |
| **theta-gamma PAC (log10)** | parahippocampal | +0.45 (0.20) | +0.74 (0.035) | -0.05 (0.91; 1.00) | -0.03 (0.92) | +0.06 (0.85) |
| **aperiodic exponent** | MTL composite | -0.02 (0.94) | -0.11 (0.80) | +0.00 (1.00; 1.00) | +0.20 (0.54) | +0.08 (0.80) |
| **aperiodic exponent** | entorhinal | -0.10 (0.94) | -0.08 (0.86) | -0.10 (0.82; 1.00) | +0.06 (0.86) | +0.14 (0.66) |
| **aperiodic exponent** | hippocampus | +0.05 (0.94) | -0.17 (0.68) | +0.00 (1.00; 1.00) | +0.30 (0.35) | +0.27 (0.40) |
| **aperiodic exponent** | amygdala | +0.03 (0.94) | -0.03 (0.95) | +0.07 (0.87; 1.00) | +0.23 (0.47) | -0.13 (0.68) |
| **aperiodic exponent** | parahippocampal | -0.11 (0.94) | -0.03 (0.95) | +0.02 (0.96; 1.00) | -0.03 (0.93) | +0.00 (1.00) |

**Figure 3.**
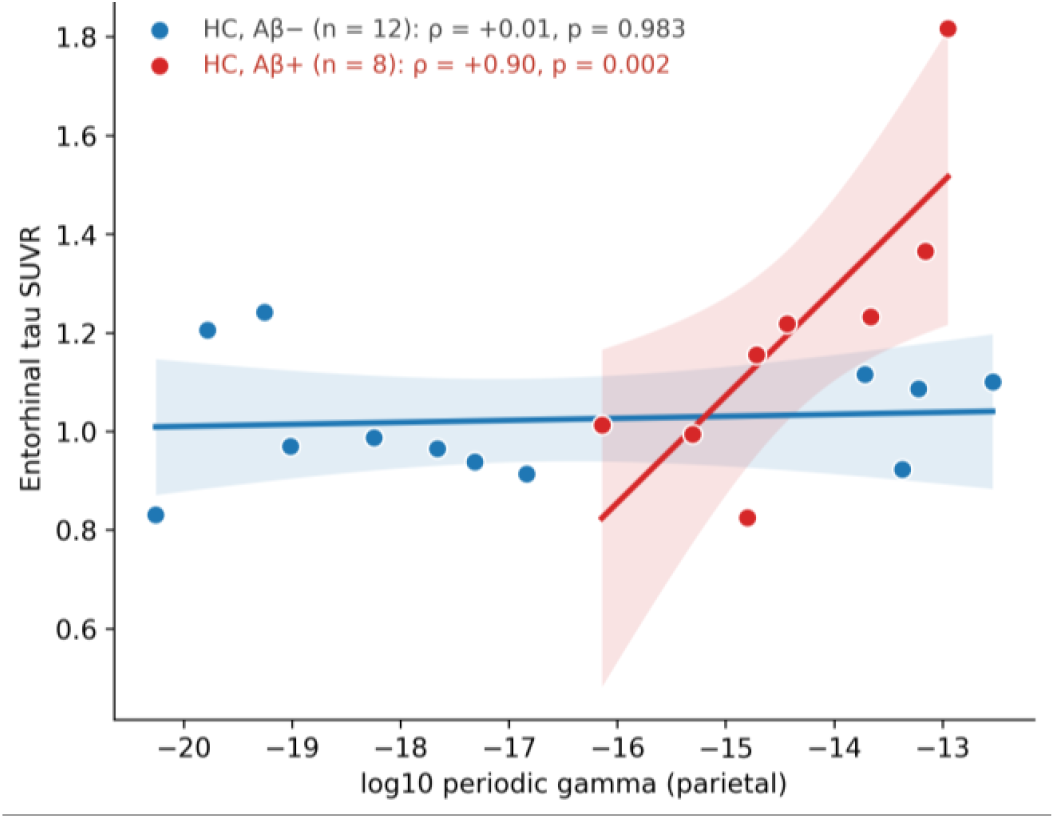
Parietal periodic gamma and MTL tau. Periodic gamma and entorhinal tau in Aβ-negative (blue) and Aβ-positive (red) HC, with separate linear fits and 95% CI. The significant association is shown in red font.

### 3.5. Parietal periodic gamma moderates the Aβ-tau relationship

Periodic gamma moderated the association between Centiloid and MTL composite tau (interaction β=+0.22, 95% CI 0.05 to 0.38, p=0.012; **Figure 4**): at higher Centiloid (+1 SD), higher periodic gamma was associated with higher MTL tau (β=+0.35, p=0.025), whereas at lower Centiloid (−1 SD), it was not (β=-0.09, p=0.084). The interaction survived FDR correction in all five MTL regions (p=0.0078-0.025, q=0.029-0.045). Theta-gamma PAC interactions also survived FDR correction in all five regions (q=0.029-0.031) but were significant after sensitivity analyses excluding one extreme subject. Alpha-gamma PAC showed a nominal interaction for the amygdala only (q=0.041), which was likewise not robust, and the aperiodic exponent showed no moderation. The moderation finding is therefore specific to periodic gamma and consistent across MTL regions.

**Figure 4.**
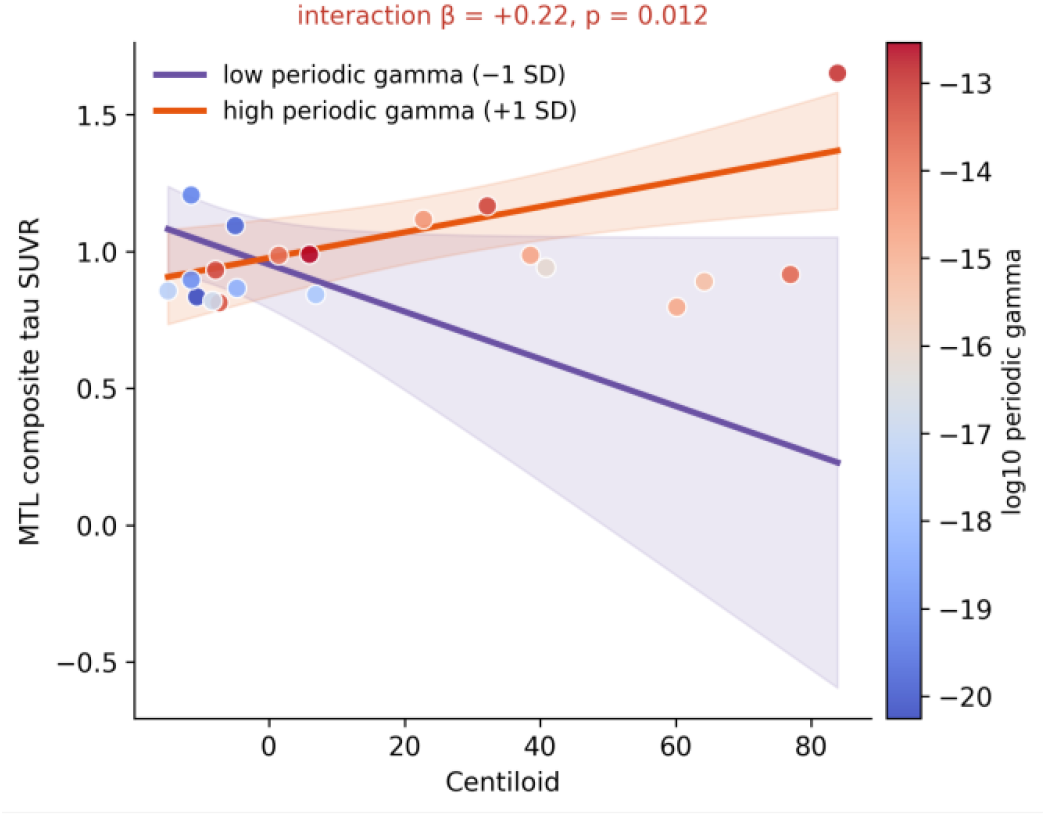
Parietal periodic gamma and MTL tau. Model-predicted MTL composite tau across Centiloid at low and high periodic gamma (20 HC with tau-PET). The significant associations are shown in red font.

### 3.6. In Aβ-positive normal individuals, parietal periodic gamma may track the tau accumulation and spread at the individual level

Finally, among the eight Aβ-positive normal individuals, tau was higher in the individuals with the highest periodic gamma and lower in the individuals with the lowest periodic gamma (**Figure 5**). The individual with the highest periodic gamma showed the most widespread tau elevation, spanning medial temporal, lateral temporal, occipital, and parietal regions, and the individual with the next highest periodic gamma showed tau elevation focused on the medial temporal lobe (entorhinal, parahippocampal, and hippocampal regions). This suggests that periodic gamma may relate to tau even at the individual level.

**Figure 5.**
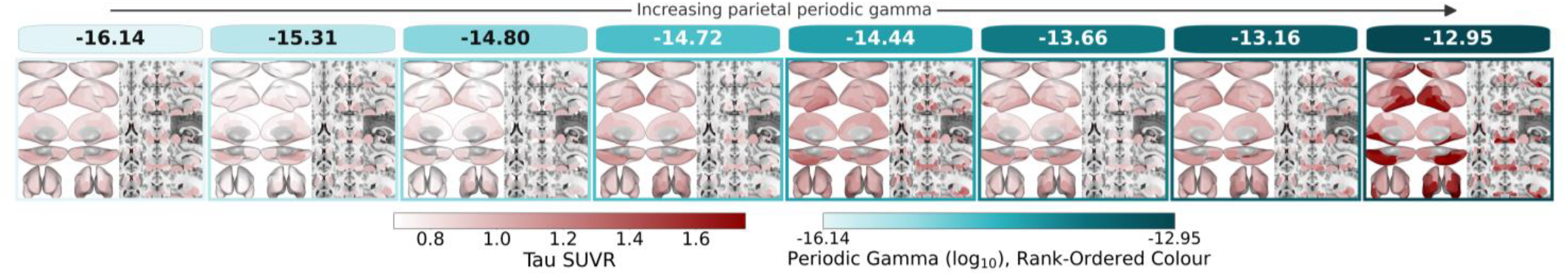
Regional tau by periodic gamma in our eight Aβ-positive normal adults (ordered from lowest to highest parietal periodic gamma).

## 4. DISCUSSION

In this pilot study, our main goal was to test whether parietal resting EEG measures of E-I balance carry information about Aβ and tau in cognitively normal older adults. We found four main results. First, higher Aβ burden was associated with higher parietal periodic gamma power in HC, consistently across robust estimators, although the association did not survive correction across the four E-I measures; the other E-I measures (alpha-gamma and theta-gamma PAC and the aperiodic exponent) were not related to Aβ. Second, E-I measures were not associated with MTL tau across all participants, but among Aβ-positive participants, higher periodic gamma was associated with higher tau in the MTL, most strongly in the entorhinal cortex, whereas no association was present in Aβ-negative participants. Third, and most importantly, periodic gamma moderated the relationship between Aβ and tau: the association between Aβ burden and MTL tau was steeper in participants with higher periodic gamma, consistently across MTL regions and specifically for periodic gamma among the E-I measures. Fourth, at the individual level, the participants with the highest periodic gamma had the greatest tau, with widespread elevation in the highest and medial temporal elevation in the next highest.

Our first finding was that higher Aβ burden was associated with higher parietal periodic gamma power in cognitively normal adults, whereas the other E-I measures (alpha-gamma and theta-gamma PAC and the aperiodic exponent) were not related to Aβ. The parietal cortex is among the earliest sites of Aβ deposition, and its high baseline activity makes it particularly vulnerable to Aβ-related changes in excitability. This pattern is broadly consistent with electromagnetic studies reporting detectable oscillatory change in preclinical and prodromal AD.^25, 26^ At the cellular level, Aβ pathology directly disrupts inhibitory circuitry by altering receptor function, modifying subunit composition, and selectively impairing parvalbumin (PV) interneurons.^27^ PV interneurons are critical for generating EEG gamma oscillations and supporting processes for memory and attention.^28^ PV interneuron dysfunction leads to disrupted oscillatory rhythms and network hyperactivity.^29-31^ In parallel, Aβ oligomers also directly impair GABAergic signaling via oxidative stress, synaptic dysfunction, and disrupted receptor trafficking^32^.

We next tested the direct EEG-tau relationship. Across all healthy controls, none of the four E-I measures were associated with MTL tau after correction for multiple comparisons. Alpha-gamma PAC showed the largest nominal associations. EEG-based gamma PAC has also been reported to be sensitive in detecting brain hyperexcitability.^33^ In contrast, among Aβ-positive participants, higher periodic gamma was associated with higher MTL tau, most strongly in the entorhinal cortex (ρ = 0.90, FDR q = 0.040), which is the first site of tau accumulation in Braak staging^34, 35^, whereas no association was present in Aβ-negative participants. This pattern suggests that cortical excitability is related to tau mainly in the presence of abnormal Aβ. This is consistent with evidence that neuronal hyperactivity increases tau release and spread in vivo^36^, and tau accumulates faster in regions functionally connected to existing tau^37^. We speculate that this relationship is mainly through an excitatory effect, as a recent study showed that this EEG power spectrum is associated with higher concentrations of the excitatory neurotransmitter, glutamate.^38^ Mechanistically, this hyperexcitability may arise from both increased glutamatergic excitation and reduced GABAergic inhibition.^29^ Over time, sustained hyperexcitability can progress to excitotoxicity^39^, aberrant network reorganization, and ultimately neuronal silencing mediated by concomitant tau accumulation ^40^and AD progression.

Our moderation result has a likely mechanistic basis. Parietal gamma rhythms depend on fast-spiking, parvalbumin-expressing inhibitory interneurons, and gamma power is therefore a candidate scalp-level index of E-I balance. In our data, periodic gamma moderated the association between Aβ burden and MTL tau: the relationship between Aβ and tau was steeper in participants with higher periodic gamma, consistently across all five MTL regions, and this effect was specific to periodic gamma among the E-I measures. Although this moderation depended on a small number of participants and requires confirmation, it is consistent with the hypothesis from our previous work^13, 14^ that excitatory neuronal responses lead to aberrant network hyperconnectivity in preclinical Aβ-positive individuals at high risk of developing AD, and that this hyperconnectivity increases the risk of tau spread and disease progression.^13, 14^ In another study, we also investigated whether there could be associations between Aβ and tau across spatially distinct regions, and we found strong remote associations between early-stage tau accumulation in the MTL and cortical Aβ across different brain regions.^41^ A study using transgenic mice with both Aβ and tau showed that Aβ-related neuronal hyperactivity plays a key role in driving tau spread, starting in the entorhinal cortex and extending to connected brain regions.^42^ In line with our central hypothesis, a recent study reported a U-shaped relationship between Aβ burden and EEG metrics in different frequency bands in individuals with evidence of neurodegeneration.^26^ EEG-based parameters, especially in the gamma frequency, are critical in establishing synchronization, not only in short-distance cortical brain connectivity,^43^ but also in long-range brain connectivity^44-46^. Higher periodic gamma power with Aβ burden may directly reflect early network hyperexcitability^39, 47^, which is specifically influenced by E-I GABAergic signaling, glutamatergic tone, and neurotransmitters ^48-50^.

Among the eight Aβ-positive cognitively normal adults, tau was highest in the individuals with the highest periodic gamma and uniformly low in the individuals whose periodic gamma was low, with the most widespread elevation in the individual with the highest value. This ordering is consistent with experimental evidence that Aβ-related network hyperexcitability promotes tau release and spread^36^ and with recent human imaging showing that Aβ-driven hyperexcitability of the default mode network precedes tau accumulation in medial temporal regions.^51^ Periodic gamma power, which depends on the activity of inhibitory interneurons, may therefore index this excitatory shift.

This study has several limitations. First, the sample was small, explanatory, and from a single site, so effect sizes are imprecise and may not generalize. Second, the design was cross-sectional. Tau was measured at one time point, so we could not test whether EEG markers predict tau accumulation or spread, and the associations with “accumulation” should be read as associations with tau burden. Third, gamma-band power is vulnerable to residual muscle and line-noise artifact despite our preprocessing, and we did not record electromyography.

In this pilot study, a short resting EEG recording carried information related to Aβ and tau burden in cognitively normal older adults. Parietal periodic gamma power, a marker of cortical excitability, increased with Aβ burden and was associated with medial temporal tau only in the presence of amyloid, most clearly in the entorhinal cortex. Periodic gamma also moderated the association between Aβ and MTL tau, whereas phase-amplitude coupling and the aperiodic exponent showed no robust relationships. Periodic gamma can therefore be considered as a candidate marker of Aβ-related E-I imbalance in preclinical AD. Because the sample was small and cross-sectional, these findings are hypothesis-generating and require confirmation in larger, longitudinal cohorts.

## Data Availability

All data produced in the present study are available upon reasonable request to the authors

## DECLARATIONS

### Ethics approval and consent to participate

The Institutional Review Board of Weill Cornell Medicine gave ethical approval for this work. All participants provided written informed consent before participation.

### Data availability

The data supporting the findings of this study are available from the corresponding author upon reasonable request.

### Code availability

The analysis code used in this study is available from the corresponding author upon reasonable request.

### Funding

This work was not supported.

### Competing interests

The authors declare no competing interests.

GLP-1RA: glucagon-like peptide 1 receptor agonist; SD: standard deviation; M: male; DM: diabetes mellitus; BMI: body mass index; ALSFRS-R: revised amyotrophic lateral sclerosis functional rating scale.

## Notes

### Competing Interest Statement

The authors have declared no competing interest.

### Author Declarations

Institutional Review Board of Weill Cornell Medicine gave ethical approval for this work

